# Facility-based abortion care in India, 2008-2022: long-run stagnation, pandemic disruption, and an early shift toward later-gestation procedures after the MTP Amendment Act

**DOI:** 10.64898/2026.09.06.26362364

**Authors:** Vartika Mishra, Koshtubh Singh Parihar

## Abstract

**Background:** Medical termination of pregnancy (MTP) has been legal in India since 1971, and the MTP (Amendment) Act 2021 expanded gestational limits and eligibility. Yet trends in facility-based abortion provision, their disruption by COVID-19, and any early response to the 2021 reform have not been described from routine national data. We quantified 14 years of public and private facility MTP provision by gestational age, geographic access, pandemic disruption, and the post-Amendment shift in later-gestation procedures.

**Methods:** Analysis of India’s Health Management Information System (HMIS), fiscal years 2008-09 to 2021-22. We extracted monthly MTP counts by gestational band (up to 12 weeks; more than 12 weeks) and sector, post-abortion care indicators, and abortion-related maternal deaths for all districts and states, harmonising definitions across the 2017-18 schema change. We described national and state trends, the share of districts reporting zero MTPs, and — for 2017-2022 — used interrupted time-series models to quantify COVID-19 disruption; we compared the more-than-12-week share before and after the MTP Amendment Act rules (September 2021).

**Results:** Reported facility MTPs were essentially flat over 14 years, at roughly 0.6-0.74 million per year, despite population growth. The more-than-12-week share of MTPs halved over the period, from 17.5% in 2008-09 to 6.9-9.5% after 2017. Around 5% of districts reported zero MTPs in any given year. COVID-19 reduced total MTPs by about 25% below expected over March 2020-March 2022, with public-sector provision hit harder (-30%) than private (-20%), and post-abortion contraception falling by half and not recovering. Strikingly, more-than-12-week MTPs surged to 155% of expected after the MTP Amendment Act rules took effect in September 2021, and the more-than-12-week share rose from 6.9% pre-pandemic to 9.7% post-Amendment. State MTP deficits over the pandemic exceeded 40% in Rajasthan, Delhi, Assam and Tamil Nadu.

**Conclusion:** Facility-based abortion provision in India has stagnated in absolute terms for over a decade and is unequally distributed, consistent with the large majority of abortions occurring outside the facility system. COVID-19 caused a substantial, public-sector-concentrated disruption from which post-abortion contraception did not recover, while the earliest post-Amendment data show a measurable shift toward later-gestation facility procedures. Routine data provide an essential, if partial, monitoring tool for abortion-care policy.

## Introduction

India legalised abortion relatively early, through the Medical Termination of Pregnancy (MTP) Act of 1971, and substantially liberalised it through the MTP (Amendment) Act 2021, which raised the upper gestational limit for specified categories from 20 to 24 weeks, extended eligibility to unmarried women, and revised provider and approval requirements [1,2]. Despite this progressive legal framework, abortion access in India is shaped less by law than by service availability: the landmark 2015 national incidence study estimated 15.6 million abortions annually, of which only around one-quarter occurred in health facilities, the majority being medication abortions obtained outside the formal system [3]. Facility-based provision, though a minority of all abortions, is the component the health system directly controls, funds and can expand — and the only component captured in routine data.

How facility-based abortion provision has evolved over time, how unequally it is distributed, how it was affected by the COVID-19 pandemic, and whether the 2021 Amendment produced any early change in practice have not been described from routine national data. India’s Health Management Information System (HMIS) records monthly MTP counts by gestational band and sector from every district. We used 14 years of these records to describe long-run trends and geographic inequality in facility MTP provision, to quantify pandemic disruption and its differential impact on public versus private provision and on post-abortion care, and to examine the earliest post-Amendment signal in later-gestation procedures. Throughout, we interpret these data as measuring facility-based, health-system-reported provision — not total abortion incidence, most of which is invisible to HMIS.

## Methods

### Data source and indicators

We used monthly district-level HMIS reports, fiscal years 2008-09 to 2021-22 (April 2008-March 2022). We extracted MTPs by gestational band (up to 12 weeks; more than 12 weeks) and by sector (public, private), harmonising the pre- and post-2017 indicator definitions; total MTPs; post-abortion/MTP complications identified and treated; post-abortion contraception provision (2017 onward); and abortion-related maternal deaths. In the pre-2017 schema, gestational bands were reported for public institutions with a separate private total; from 2017-18, both bands are reported with public and private sub-columns, so the gestational split becomes available for both sectors — a definitional break we mark explicitly and do not splice.

### Statistical analysis

We described national and state annual MTP volumes by gestational band and sector, and the share of districts reporting zero MTPs each year as an access measure. As an internal extraction check we compared HMIS state MTP totals against the Ministry of Health’s published state-wise MTP statistics for 2010-11 to 2014-15; these matched exactly (the published series being HMIS-derived), confirming faithful extraction but providing no independent validation. For the COVID-19 analysis, restricted to the internally consistent 2017-18 to 2021-22 era, we fitted interrupted time-series models (log counts on linear time and calendar-month indicators, fitted April 2017-February 2020) and summarised observed/expected ratios by pandemic period and cumulative deficits over March 2020-March 2022, with a same-month-mean counterfactual as sensitivity. To assess the MTP Amendment Act, whose rules were notified in September 2021, we compared the more-than-12-week share of MTPs across three windows: pre-pandemic (April 2019-February 2020), the pandemic pre-Amendment period (March 2020-August 2021), and the post-Amendment period (September 2021-March 2022). Analyses used Python 3.13; reporting follows RECORD guidance [4].

## Results

### Fourteen years of stagnation and inequality

Reported facility MTPs were remarkably flat over the study period, fluctuating around 0.6-0.74 million per year with no sustained growth despite a rising population of reproductive age (Table 1, Figure 1). Within this stable total, the more-than-12-week share of MTPs fell markedly, from 17.5% in 2008-09 to 12.8% by 2010-11 and to 6.9-9.5% across the post-2017 years — a shift toward earlier-gestation procedures, consistent with expanding early medical abortion within facilities. Geographic access remained incomplete throughout: around 5% of districts reported zero MTPs in any given year (9.7% in 2008-09, improving to 4.3-5.3% subsequently), indicating persistent local access gaps.

**Table 1.** National annual facility MTPs by gestational band, more-than-12-week share, and share of districts reporting zero MTPs, 2008-09 to 2021-22.

| Fiscal year | Total MTPs | ≤12 weeks | >12 weeks | >12-week share (%) | Zero-MTP districts (%) |
| --- | --- | --- | --- | --- | --- |
| 2008-2009 | 629,327 | 441,347 | 93,668 | 17.5 | 9.7 |
| 2009-2010 | 648,587 | 392,998 | 64,976 | 14.2 | 5.4 |
| 2010-2011 | 642,305 | 398,616 | 58,357 | 12.8 | 5.4 |
| 2011-2012 | 620,241 | 319,174 | 45,458 | 12.5 | 4.6 |
| 2012-2013 | 639,884 | 316,888 | 35,964 | 10.2 | 5.3 |
| 2013-2014 | 665,789 | 341,068 | 45,475 | 11.8 | 5.1 |
| 2014-2015 | 700,358 | 351,430 | 42,692 | 10.8 | 4.9 |
| 2015-2016 | 720,835 | 352,668 | 42,281 | 10.7 | 5.3 |
| 2016-2017 | 742,486 | 347,522 | 44,565 | 11.4 | 5.3 |
| 2017-2018 | 696,569 | 633,418 | 63,151 | 9.1 | 4.5 |
| 2018-2019 | 725,483 | 673,820 | 51,663 | 7.1 | 4.7 |
| 2019-2020 | 715,087 | 665,637 | 49,450 | 6.9 | 4.3 |
| 2020-2021 | 535,671 | 493,875 | 41,796 | 7.8 | 6.0 |
| 2021-2022 | 595,049 | 538,688 | 56,361 | 9.5 | 5.1 |

**Figure 1.**
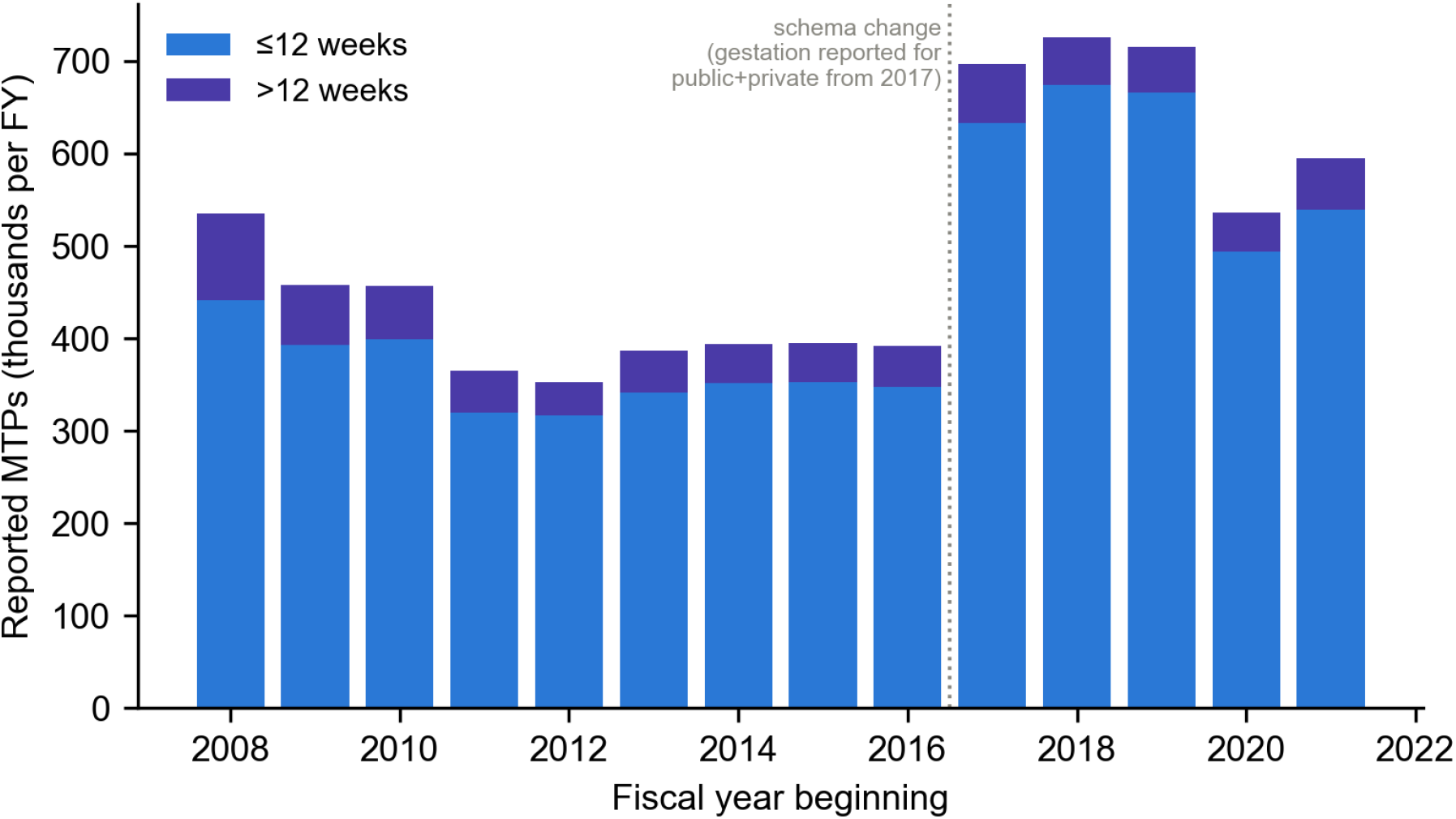
National annual facility MTPs by gestational band, 2008-2022; dotted line marks the 2017-18 schema change after which gestational bands are reported for both sectors.

As an extraction check, HMIS state MTP totals for 2010-11 to 2014-15 matched the Ministry of Health’s published state-wise MTP statistics exactly, confirming that the published national statistics are themselves HMIS-derived; this validates our extraction but means the two sources are not independent.

### Pandemic disruption

COVID-19 substantially reduced facility MTP provision (Table 2, Figure 2). Total MTPs fell to 62% of expected in April 2020 and remained depressed, with a cumulative deficit of 22-25% over March 2020-March 2022 across counterfactuals — on the order of several hundred thousand fewer facility abortions. Public-sector provision was hit harder (cumulative deficit ∼30%) than private (∼20%), suggesting some shift toward private facilities during the pandemic. Post-abortion contraception was the most severely and persistently affected indicator, falling to 47% of expected in April 2020 and remaining around half of expected through the entire period — a durable gap in the post-abortion care cascade. State MTP deficits were highly uneven, exceeding 40% in Rajasthan, Delhi, Assam and Tamil Nadu (Table 3, Figure 3).

**Table 2.**
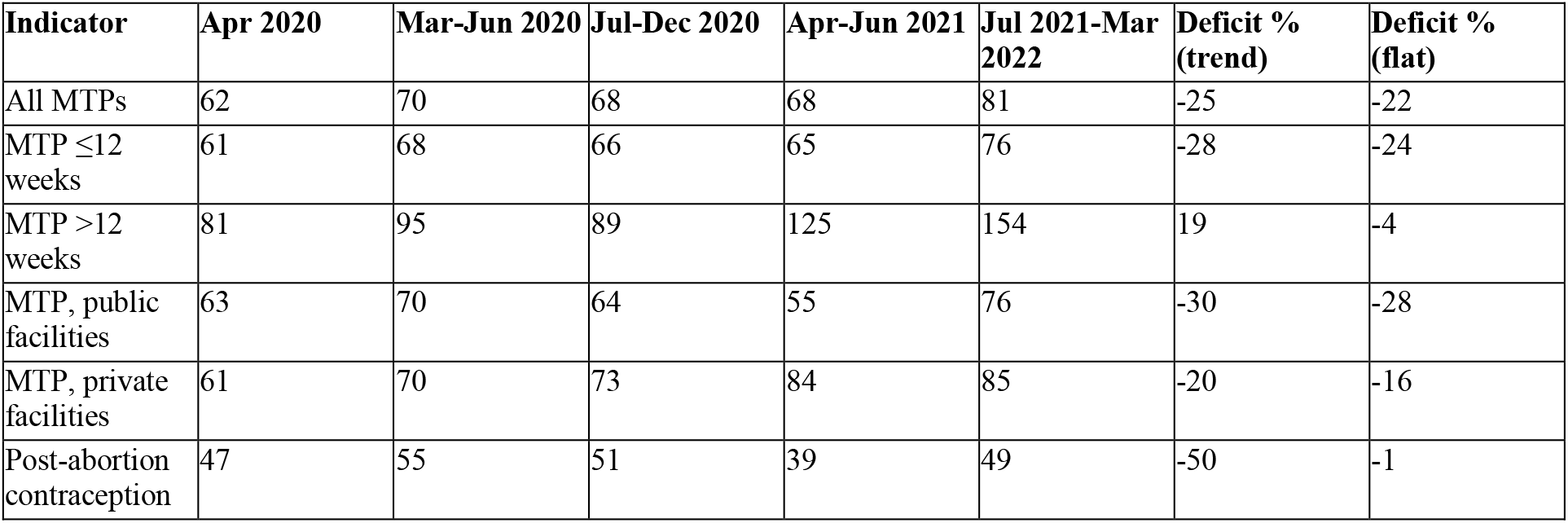
National observed/expected ratios (%) by pandemic period and cumulative deficits (March 2020-March 2022) under two counterfactuals, MTP and post-abortion care indicators.

**Table 3.** State cumulative MTP deficits, March 2020-March 2022 (states with >10,000 expected MTPs; thousands).

| State | Observed (000s) | Expected (000s) | Deficit (%) |
| --- | --- | --- | --- |
| Rajasthan | 43.8 | 84.4 | -48.1 |
| Delhi | 23.3 | 42.1 | -44.8 |
| Assam | 134.1 | 229.4 | -41.6 |
| Tamil Nadu | 174.9 | 286.6 | -39.0 |
| Uttar Pradesh | 56.4 | 89.6 | -37.0 |
| Himachal Pradesh | 7.6 | 11.6 | -34.0 |
| Chhattisgarh | 18.3 | 27.7 | -33.8 |
| West Bengal | 79.1 | 115.9 | -31.8 |
| Punjab | 45.2 | 63.3 | -28.6 |
| Gujarat | 20.3 | 28.3 | -28.0 |
| Jharkhand | 12.8 | 17.6 | -27.3 |
| Karnataka | 58.6 | 80.4 | -27.2 |
| Kerala | 20.3 | 27.1 | -25.0 |
| Madhya Pradesh | 69.6 | 78.6 | -11.5 |
| Haryana | 69.6 | 78.6 | -11.5 |
| Maharashtra | 255.1 | 286.7 | -11.0 |
| Odisha | 25.1 | 28.1 | -10.6 |
| Jammu & Kashmir | 9.1 | 10.1 | -9.7 |
| Bihar | 16.3 | 14.9 | 9.7 |

**Figure 2.**
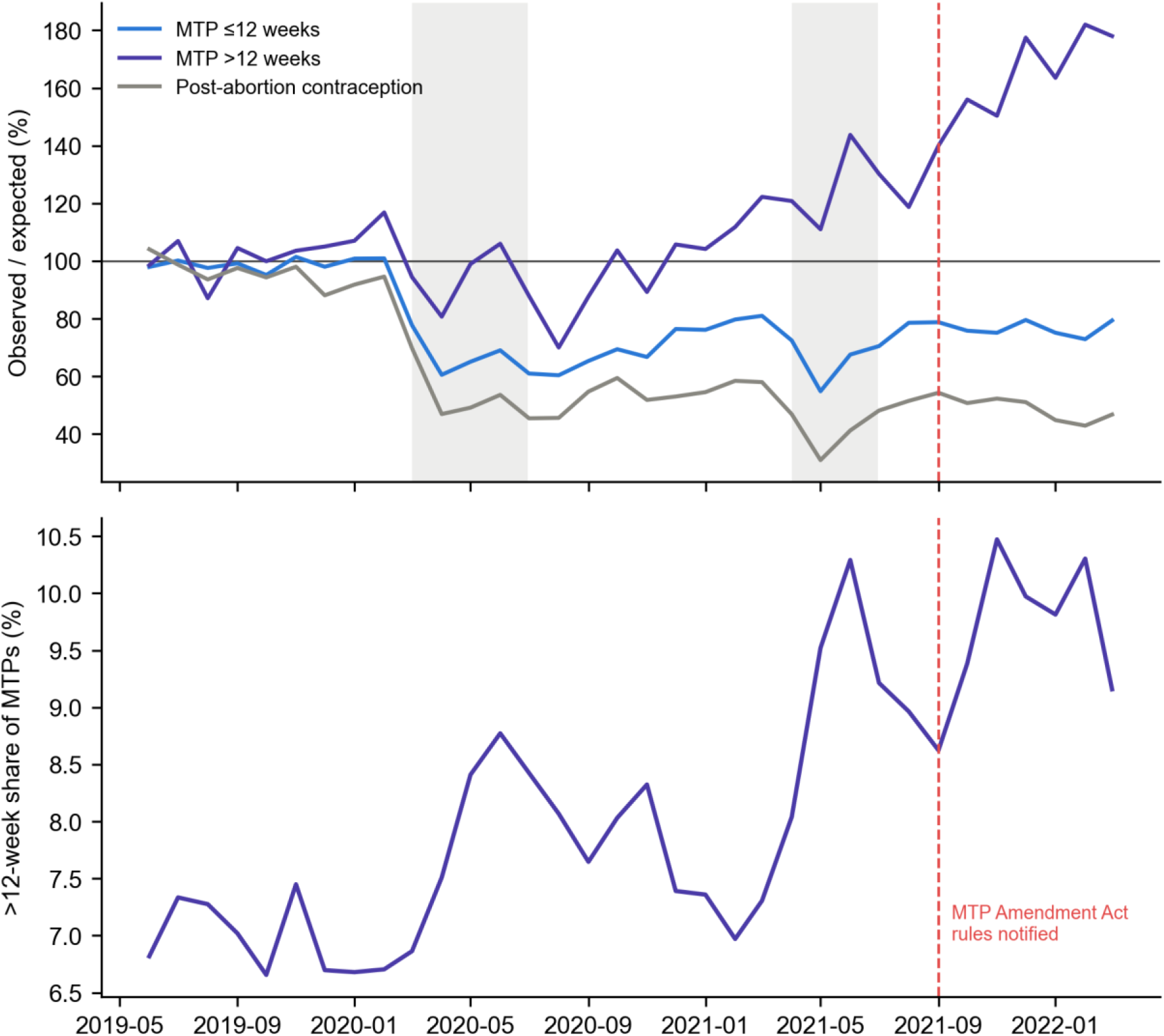
Observed/expected ratios for MTPs by gestational band and post-abortion contraception across the two COVID-19 waves (top), and the more-than-12-week share of MTPs (bottom); dashed line marks notification of the MTP Amendment Act rules (September 2021).

**Figure 3.**
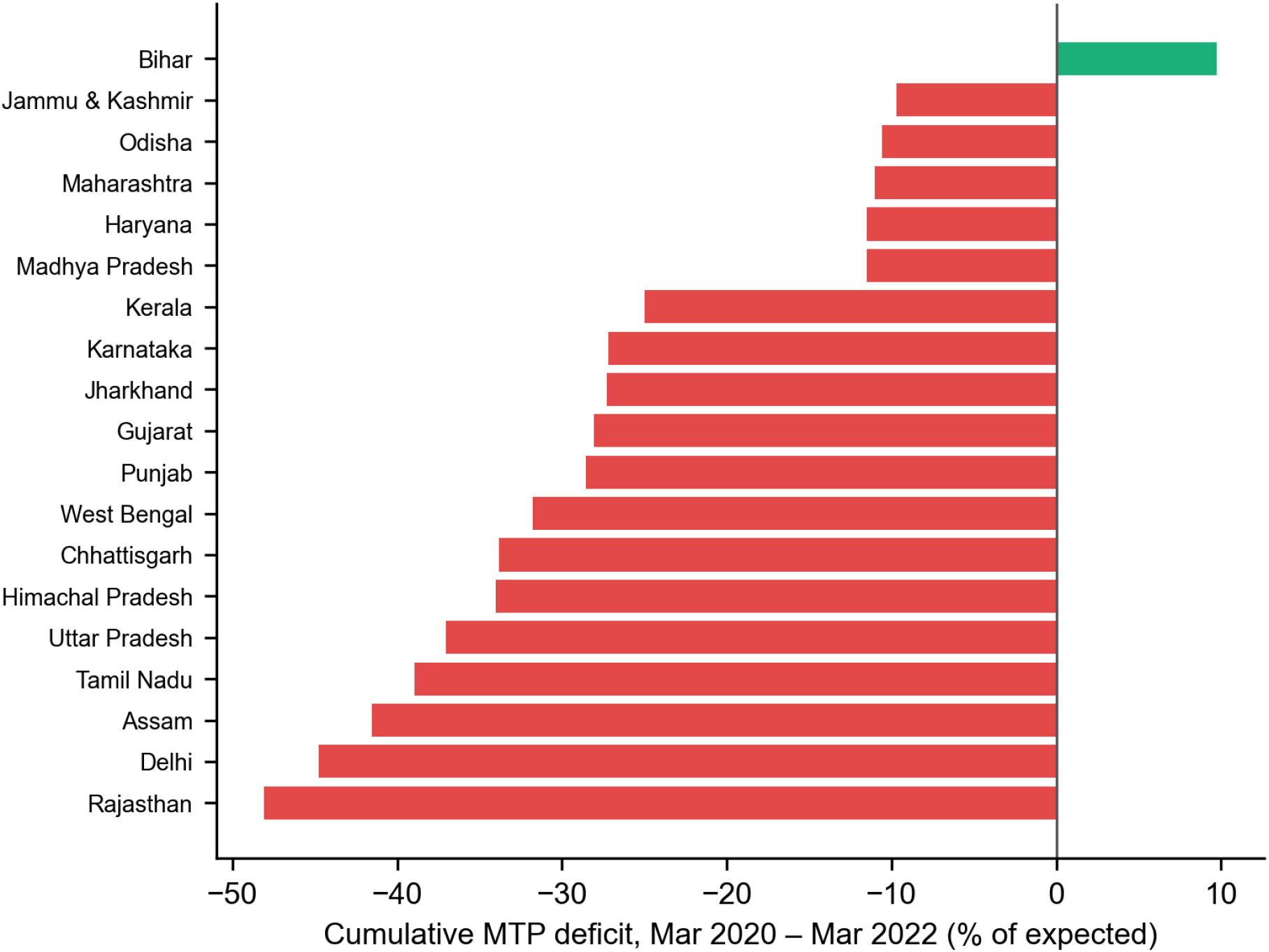
State cumulative MTP deficit, March 2020-March 2022 (% of expected).

### An early post-Amendment shift toward later-gestation procedures

The more-than-12-week MTP series showed a distinctive pattern diverging from the up-to-12-week series (Figure 2). While up-to-12-week MTPs remained around 75-80% of expected through the pandemic, more-than-12-week procedures rose above expected from mid-2021 and surged to 155% of expected in the July 2021-March 2022 period, immediately following the notification of the MTP Amendment Act rules in September 2021. Correspondingly, the more-than-12-week share of MTPs rose from 6.9% pre-pandemic to 8.2% during the pandemic pre-Amendment window and to 9.7% post-Amendment. This early signal is consistent with the Amendment’s expansion of the upper gestational limit enabling more later-gestation procedures within the facility system, although the short post-Amendment window (seven months) warrants cautious interpretation and longer follow-up.

## Discussion

Fourteen years of routine data show that facility-based abortion provision in India has stagnated in absolute terms, is unequally distributed across districts, was materially disrupted by COVID-19, and shows an early upturn in later-gestation procedures following the 2021 Amendment. The flat facility total, against a backdrop of an estimated 15.6 million annual abortions nationally [3], underlines that the facility system handles a minority of abortions and has not expanded its share — most abortion care continues to occur through medication abortion outside routine reporting, a structural feature that any facility-based monitoring, including ours, cannot capture.

The pandemic findings carry specific implications. The concentration of losses in the public sector, and the collapse of post-abortion contraception to half its expected level with no recovery, identify the post-abortion care cascade as a fragile link that health-system shocks readily break. Because post-abortion contraception is a proven lever against repeat unintended pregnancy, its durable deficit is a concrete, addressable target for service recovery. The harder hit to public provision also raises equity concerns, since public facilities disproportionately serve women who cannot afford private care.

The post-Amendment shift toward later-gestation facility procedures is, to our knowledge, the first quantitative signal of the 2021 Act’s effect on practice. It is biologically and legally coherent — the Amendment specifically raised the gestational ceiling — and appears as a clear divergence between gestational bands rather than a general trend. Nonetheless, the window is short, the change is measured against a pandemic-perturbed baseline, and later-gestation counts are small; we therefore frame it as an early signal for continued monitoring rather than a settled effect. The interrupted-time-series divergence between bands, however, is difficult to attribute to reporting artefact, which would not selectively elevate one gestational band.

### Strengths and limitations

Strengths include a continuous 14-year national series at monthly resolution, gestational and sector detail, district-level access measurement, dual counterfactuals, and a policy-aligned pre/post-Amendment comparison. Limitations are fundamental to the data source and we stress them: HMIS captures facility-reported, predominantly public-sector abortion provision only, and is blind to the majority of abortions that occur as medication abortion outside facilities, so our counts must not be read as abortion incidence; private-sector reporting into HMIS is incomplete, particularly in early years, so sector comparisons and the apparent public-private pandemic divergence should be interpreted cautiously; the 2017-18 schema change alters how gestational bands are captured by sector, so we avoid cross-era splicing of the sector-specific series; the post-Amendment window is brief; and abortion-related maternal-death counts are small and under-reported. Finally, service counts cannot address abortion safety, client experience, or the large informal sector.

### Conclusion

Facility-based abortion care in India has been static and unequal for over a decade, was disrupted by COVID-19 with a lasting hit to post-abortion contraception, and shows an early post-Amendment shift toward later-gestation procedures. Routine data are an essential, if inherently partial, tool for monitoring abortion-care policy, and point to post-abortion contraception recovery and geographic access as immediate priorities.

## Data Availability

All data produced in the present study are available upon reasonable request to the authors.

https://hmis.mohfw.gov.in/#!/standardReports

https://data.gov.in/

